# Virtual care with digital technologies for rural and remote Canadians living with cardiovascular disease

**DOI:** 10.1101/2020.12.17.20248333

**Authors:** Ryan Buyting, Sarah Melville, Hanif Chatur, Christopher W. White, Jean-François Légaré, Sohrab Lutchmedial, Keith R. Brunt

## Abstract

Canada is a wealthy nation with a geographically diverse population, seeking health innovations to better serve patients in accordance with the Canada Health Act. In this country, population and geography converge with social determinants, policy, procurement regulations, and technological advances, in order to achieve equity in the management and distribution of healthcare. Rural and remote patients are a vulnerable population; when managing chronic conditions such as cardiovascular disease, there is inequity when it comes to accessing specialist physicians at the recommended frequency—increasing the likelihood of poor health outcomes. Ensuring equitable care for this population is an unrealized priority of several provincial and federal government mandates. Virtual care technology may provide practical, economical, and innovative solutions to remedy this discrepancy. Here we review the literature pertaining to the use of virtual care technologies to monitor patients with cardiovascular disease living in rural areas of Canada. A search strategy was developed to identify the literature specific to this context across three bibliographic databases. 166 unique citations were ultimately assessed for eligibility, of which 36 met the inclusion criteria. In our assessment of these articles, we provide a summary of the interventions studied, their reported effectiveness in reducing adverse events and mortality, the challenges to implementation, and the receptivity of these technologies amongst patients, providers and policy makers. Further, we glean insight into the barriers and opportunities to ensure equitable care for rural patients and conclude that there is an ongoing need for clinical trials assessing virtual care technologies in this context.

**Summary:** Patients living in rural and remote communities’ experience diverse challenges to receiving equitable healthcare as is mandated by the Canada Health Act. Advances in virtual care technology may provide practical, economical, and innovative solutions to ensure this for patients in remote and rural living situations. Here we provide a state-of-the-art review of virtual care technologies available to patients with cardiovascular disease living in rural areas of Canada.

## Introduction

Based on its geographic size and distributed population density, Canada is justified to become a leader of innovation for remote medical care—whether the nation is optimally capitalizing on this is unclear. Rural populations are essential to the well-being of a nation, as they are naturalized environmental stewards; farmers and harvesters providing the majority of food, and other natural resources that enable urban living and a trade economy. In contrast to urban populations, rural patients are a vulnerable group in need of special attention to ensure equitable healthcare delivery. ^1-3^ The inequities impacting rural populations are complex, but a major concern is timely access to specialist physicians in accordance with clinical guideline recommendations. ^4-6^ Disparities in healthcare negatively impact all clinical outcomes, and thus provisions to secure equitable healthcare and comparable health outcomes for vulnerable groups are necessary. The scientific and clinical efforts toward addressing this are increasing and multiple approaches to democratize care are being studied. ^7^ One approach is the opportunity for healthcare workers, researchers, and policymakers to adopt innovative virtual care and remote-monitoring platforms to improve clinical outcomes, reduce hospital wait times, or increase the quality of life for patients and their families. ^8, 9^ An example of how such a system could work is depicted in Figure 1, which has been adapted from Prescher *et al*. ^10^ In the 1970s, Canada was a leader in virtual care, yet now lags behind other countries. There is increasing demand by Canadian physicians for more virtual care technology, training, and payment models to support and integrate with standard clinical practice. In an organized initiative, the Canadian Medical Association (CMA) Virtual Care Task Force recently created a national strategy of practice recommendations. ^11^

**Figure 1.**
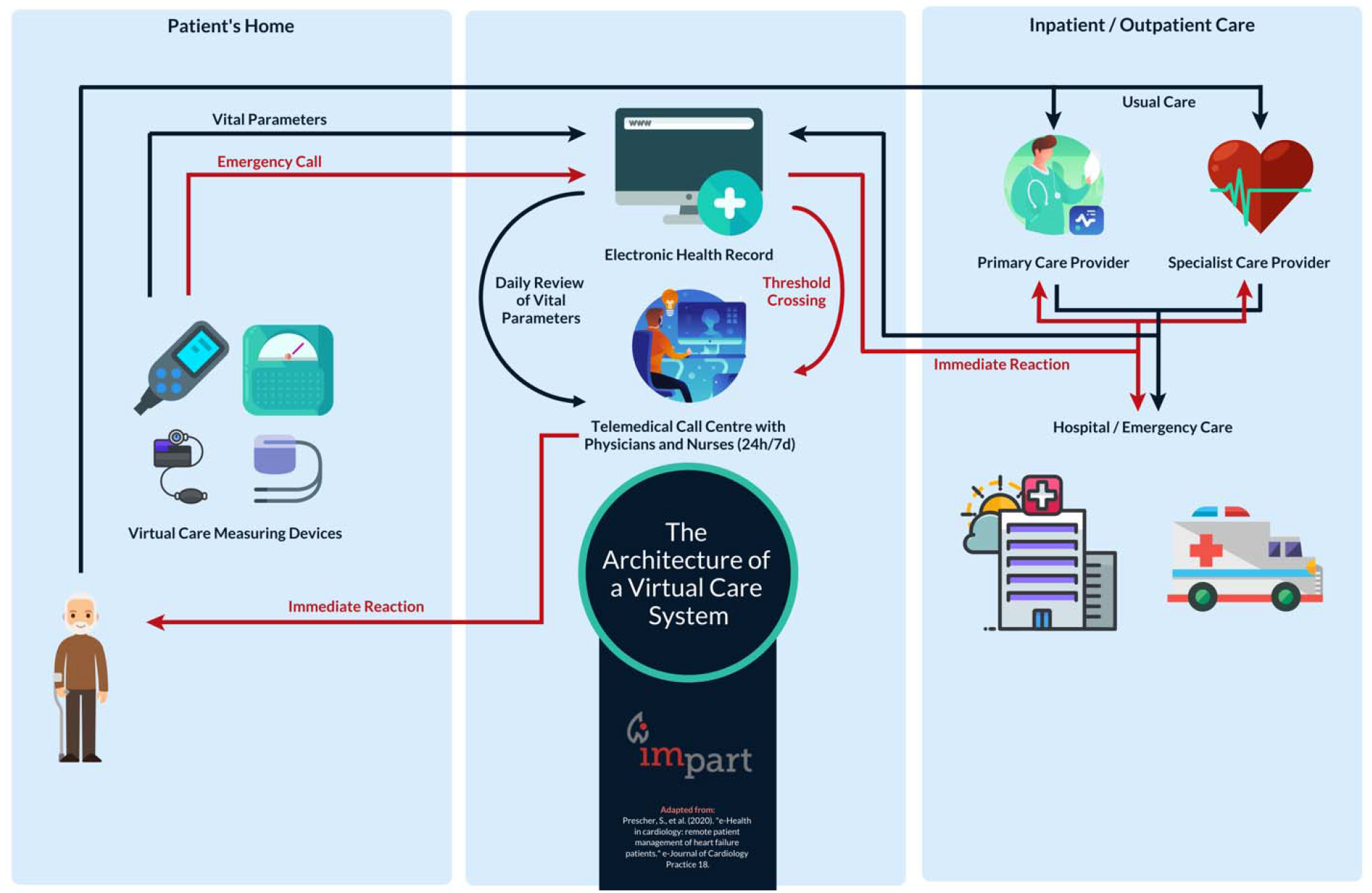
The architecture of a virtual care system.

There is no widely accepted definition of virtual care, telehealth, telemedicine, or e-medicine, and these terms are often used interchangeably, along with many others. The World Health Organization defines e-health as “the cost-effective and secure use of information and communication technologies in support of health and health-related fields, including healthcare services, health surveillance, health literature, and health education, knowledge and research.” ^12^ The use of virtual care technologies continues to grow and includes electronic virtual visits, referrals, consults, prescriptions, medical-records, vital sign monitoring, digital therapeutics, care-flow ordered checklists, telepresence and robotic surgery. For the purpose of this review, virtual care is defined as healthcare services provided directly to patients using information telecommunications technology (that is, medical care provided without traditional dependence on physical contact, paper, or archaic technologies such as fax machines). We will refer to all further instances of these terms simply as virtual care. Healthcare providers can use virtual care to remotely diagnose, treat, and manage patients, in many but not all instances. Thus, virtual care ensures continuity and integration with practice level operations, not displacement or replacement. Patients can be seen virtually by primary care providers in their homes, places of work, or anywhere else they have access to the internet. The ability to connect providers and patients without regard to their respective locations is a compelling benefit of virtual care services for both patients and providers. ^13^ It is also considered to be essential infrastructure to occupational health, safety and productivity by employers by reducing down-time, sick-days and an optimized work culture. Virtual care simply provided by video and audio alone has diverse applications, but numerous disease-specific platforms and devices have also been developed to remotely provide the consulting physician with evermore information to improve decision making. These devices include stethoscopes, glucometers, oxygen saturation monitors, weigh scales, temperature, and blood pressure devices, along with more invasive technologies such as otoscopes, continuous positive airway pressure (CPAP) sleep devices, or even implantable cardiac-defibrillators. ^14, 15^

As was alluded to above, virtual care technologies are not intended to replace traditional healthcare delivery methods, but rather to augment and integrate with services to ensure more productive, efficient, and equitable healthcare for all citizens. Regular health monitoring of patients at-risk of- or diagnosed with-cardiovascular disease (CVD), has the potential to reduce the need for significant treatment interventions, including invasive cardiac surgery or readmissions for past interventions. ^16-18^ Virtual care mitigates the need for rurally located patients to defer care or make burdensome commutes to visit with specialist clinicians for follow-up appointments. This can improve post-operative health outcomes. Owing to the fact that these technologies enable more productive, efficient, sustainable, supported, and convenient care management, they have important economic implications to healthcare payers, whether they are government or private allied insurers. CVD ranks among the largest chronic comorbidity and cause of morbidity and mortality in Canada. ^19^ Therefore, our research question was directed at understanding: what efforts in the realm of rural CVD management using virtual care technologies have recently been conducted in Canada? The purpose of this review is to summarize the relevant studies, the effectiveness of virtual care systems to reduce adverse events, improve adherence to medication and lifestyle modifications to effect all-cause mortality—recognizing barriers to implementation or acceptance of these technologies among payers, patients, and providers.

## Methods

### Eligibility Criteria

Using *a priori* developed selection criteria, we included studies reporting on the use of virtual care technologies in the context of patients in rural areas of Canada living with CVD. We used the definition of CVD outlined by the World Health Organization’s International Classification of Diseases Tenth Edition (ICD-10), which encompasses all diseases of the circulatory system, primarily: 1) coronary artery disease, or ischemic heart disease, caused by low myocardial perfusion, and thus precipitating angina, myocardial infarction, and/or heart failure. 2) cerebrovascular disease, including disorders that affect the blood supply to the brain, including stroke and transient ischemic attack, and 3) peripheral artery disease, involving the limbs or claudication in peripheral vasculature. Additionally, hypertension, valvular pathologies, aortic aneurysms, and cardiac arrhythmias were all considered to be cardiovascular pathologies that fit this definition. ^20^

This review includes all study designs, including quantitative pilot and retrospective comparison studies, and qualitative analyses of the various aspects involved with the adoption of virtual care technologies by healthcare providers, patients, and their families. Therefore, the population of interest included patients diagnosed with CVD or individuals caring for patients diagnosed with CVD at the time of study. We only included studies that explicitly mentioned rurally located patients living in Canada, as the economic and political climate of a particular healthcare network has proven to be a critical factor regulating how these virtual care technologies are being disseminated and studied. Despite this, in some subsections, we reflect on Canadian studies that do not explicitly focus on rural patients in order to contextualize the state of the technology for a given disease. The articles also had to have been published in English within the last 10 years, due to the fact that the technology is rapidly evolving, and older technologies are becoming irrelevant.

### Literature Search Strategy

Literature search strategies were developed with the assistance of an experienced information services librarian (JP) who reviewed and refined the search criteria through iterative discussion. We adopted relevant elements of the Preferred Reporting Items for Systematic Reviews and Meta-Analyses (PRISMA). ^21^ A search of the literature was performed by the primary author (RB) in May of 2020 using PubMed ©, Embase ®, and the Cochrane Central Register of Controlled Trials (CENTRAL) ©. All articles that listed in the title or abstract the terms “telehealth,” “omnicare,” “e-medicine,” “electronic medicine,” “remote consultation,” or “telemedicine” were linked through the Boolean operator “and” to rural-patient-identifying terms (“rural,” “remote,” “distance,” “rural population,” and “rural health services”), as well as the terms “Canada,” and terms relating to the diseases of interest (“hypertension” or “cardiovascular disease”). MeSH terms were used where appropriate and when possible. Using the features built into each of these respective databases, the results were limited to studies published in English within the last 10 years before being imported into Covidence ©.

### Study Selection

Upon being imported into the review software, any duplicate articles were automatically removed. The remaining titles and abstracts were screened for relevance to the research question by the primary author (RB). Articles were removed if they did not meet the *a priori* defined criteria. The full texts of each of the remaining articles were then individually obtained and reviewed in-depth to determine their applicability. At this stage, each article was either removed, or included and subsequently categorized based on the subtype of CVD in the study. A miscellaneous category was created for studies that included multiple CVD subtypes. A total of 166 citations were identified and assessed for eligibility, yet only 36 studies met the inclusion criteria and are discussed in this review. (Figure 2)

**Figure 2.**
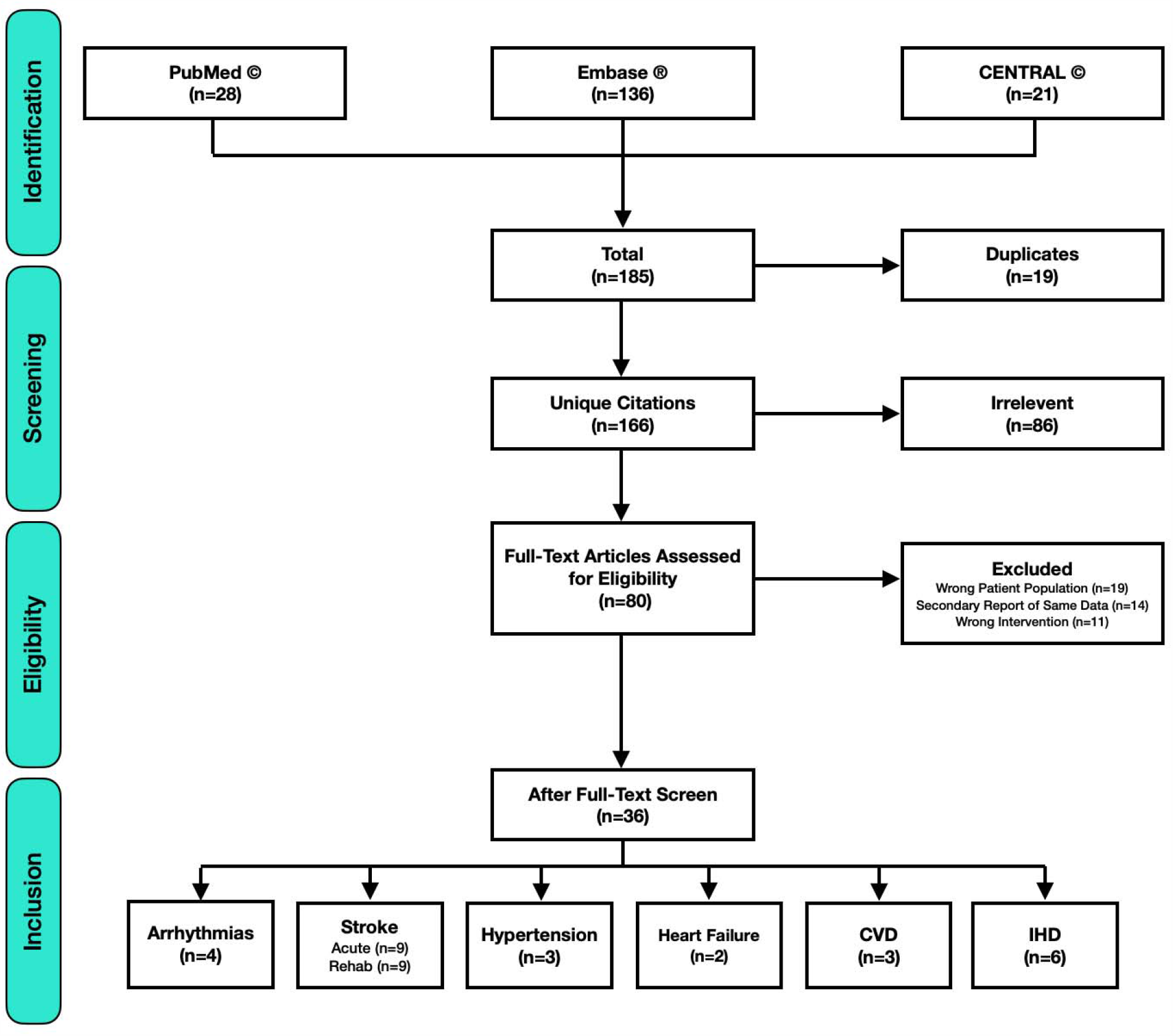
Flow chart of literature search and study extraction process, where n represents the number of studies in each group.

## Results

### Study Characteristics

The studies that met the inclusion criteria of this review discussed virtual care programs from five provinces: Ontario (15), British Columbia (10), Alberta (8), Quebec (2), and Saskatchewan (1). One study specifically included Indigenous Nations in three provinces (ON, QC, & NB), which strengthens the validity of inclusiveness in a rural Canadian context for that study. The Western Prairies (AB, SK, MB), Atlantic Canada (NB, NS, PE, NL), and Territories (YT, NT, NU) are discussed collectively due in part to collective geographic rural regionality. The article types include: 2 review articles, 1 commentary article, 1 protocol paper, and 32 original research articles (15 abstracts and 17 full-length manuscripts). The articles were published from 2010 to 2019, with more than a third (n=13) published in 2018 (n=6) and 2019 (n=7). The total number of participants in all of the studies combined was 4,014.

### General Cardiovascular Disease Interventions

Jarvis-Selinger *et al*. interviewed 48 stakeholders who were either patients diagnosed with CVD or involved in the care of patients diagnosed with CVD from rural and urban areas of British Columbia. Their goal was to better understand the role of virtual care in CVD management, the challenges to its adoption, and incentives that may increase its use among this population. The findings suggest that both healthcare providers and patients supported the use of virtual care, with the greatest benefits derived from the ability to share patient data and support patient self-management. Two concerns commonly expressed were the accuracy of patient self-reported data and security, therefore the authors state that support for implementing such systems needs to be tempered by a clear understanding of how these concerns will be mitigated. ^22^ Some reassurance may be derived from the fact that many of these systems include automatic vital sign reporting to the provider (as well as an option for manual input by the patient), and most devices adhere to federal regulatory standards with respect to privacy and security. Further support for the role of virtual care among rural patients comes from Cameron *et al*., who trialed a chronic disease self-management program in 13 rural and remote communities in Northern Ontario over a 9-month period. A total of 213 individuals with diagnoses of chronic lung disease, CVD, stroke, or arthritis were included in the study. While their goal was to better understand the psychological mechanisms underlying the effectiveness of the program itself (rather than its delivery), the fact that it was successfully conducted in this manner alone is encouraging. Specifically, the results of this study show that virtual care initiatives can be a viable means of overcoming geographic barriers for rural patients, and that patients and providers in rural areas may be uniquely receptive to these innovations. Outcomes produced from these programs, including those related to self-efficacy, mental- and physical-health status, can be comparable to in-person programs, thereby enhancing the overall reach and equity of healthcare services. ^23^ Other evidence shows that virtual care technologies used with regular exercise can improve patient cardiometabolic risk profiles; Stuckey and colleagues investigated the effects of an exercise prescription alone compared with a program supported by virtual care technology to improve cardiometabolic risk factors in rural community-dwelling adults. This work, however, seems to be ongoing, as only the rationale for this project could be sourced. ^24^ It is generally considered to be more effective to actively engage and support the person or population in establishing lifestyle changes, and with the aid of virtual care, this coaching could be delivered faster and at scale. However, more robust evidence from such programs (be it public or private) are needed to drive policy or practice changes.

### Heart Failure Interventions

Andrikopoulou and colleagues described a conceptual model of heart failure (HF) disease management, in which they state that virtual care is effective because it allows for early detection of worsening HF symptoms. Further, device data may prompt providers to optimize medication regimens and may allow patients to better understand their unique causes of decompensation. ^25^ In their 2012 article, Grant *et al*. describe the first Canadian experience using virtual care for a small sample of 25 HF patients, in which they found similar benefits, resulting in a reduced rate of all-cause patient admissions and no admissions for HF decompensation. They stressed that the success of this program was attributable to the cardiovascular nurses’ ability to assess and manage these complex patients and are therefore a key factor in the effectiveness of this model of care. ^26, 27^ Toback *et al*. later agreed with this sentiment, stating that effective virtual nursing can improve patient education and self-management, including management of worsening symptoms, daily weight measurements, medications and adjustments, appointment attendance, diet restrictions, and appropriate behaviours based on symptoms and weight changes. ^28^

Telehealth for Emergency-Community Continuity of Care Connectivity via Home Telemonitoring (TEC4Home) is a 4-year long initiative throughout many urban and rural centres in British Columbia that aims to determine how virtual care may improve care, increase patient safety during the transition of care, and determine how it is best implemented to support patients with HF. ^29^ Though still ongoing, results from their intermediary pilot study indicate that home monitoring of HF patients decreased emergency department visits and improved the overall patient experience. The length of stay data presented in the abstract suggests that patients could be discharged from the emergency department with home monitoring in order to reduce hospitalization. ^30^ Given these preliminary results, we look forward to the pending release of a peer-reviewed publication containing the results of this study.

Jaana and Sherrand assessed the comparative utilization of virtual care among patients with HF in rural versus urban environments. They describe the necessity of considering the context in which these interventions take place, which varies based on the geographic location of the patients, as manifested by a variation in the social environment, individual resources and capabilities, and the need for technology; this last point was specifically defined as a situation where virtual care can substitute for services that have disappeared or supplement existing services in a way that rural residents perceive as useful. ^31, 32^ This group conducted a cross-sectional study of all patients enrolled in the virtual care program at the University of Ottawa Heart Institute. They assessed variables associated with both the processes and outcomes of care, including the frequency in which: a patient’s vital signs were out of normal range, calls were made to a patient by the nurse, emergency department visits, hospital admissions, etc. Interestingly, there was no variation in the process or outcome measures, thus concluding that rural patients may not be perceived as extensive users of resources, nor patients who represent challenges in terms of feasibility of virtual care use. ^31^ This is encouraging, as it suggests that rural patients, who have the potential to benefit more from virtual care services than their urban counterparts, should not feel that they are being perceived as ‘stealing healthcare dollars’ from urban taxpayers when advocating for its increased promotion and use within rural communities.

### Cardiac Arrhythmia Interventions

Current practice guidelines for the treatment of cardiac arrhythmias in Canada recommend that patients with cardiovascular implantable electronic devices (CIED), undergo follow-up assessments at regular intervals. ^33-35^ Herein lies an opportunity for innovation and optimization through the implementation of virtual care technology. Amelio and colleagues describe one of the initial Canadian virtual care arrhythmia clinic experiences, which aims to improve access to electrophysiologists in Alberta. Patients are seen through a half-day clinic, scheduled on a monthly basis, by both a registered nurse and an electrophysiologist. Through a video-conference interview combined with diagnostic data supplied from the distant site, they develop a diagnosis and provide treatment options, with on-going follow-up by a nurse to ensure patients are properly managed. Patients reported satisfaction with this system and indicated saving up to $500 CAD in expenses and over 500 kilometres in travel for each appointment. ^36^

In 2013, the Canadian Cardiovascular Society and Canadian Heart Rhythm Society released a position statement regarding the use of virtual care technology for CIED follow-up. It reported that integrating these technologies into clinical practice can accelerate the identification of clinical events and device problems, therefore recommending that virtual care be available at all CIED follow-up clinics as standard of care. The report also provides advice for the proper design, implementation, and integration of a virtual care system, albeit not unique to rural populations. ^35^

Saint Paul’s Hospital in Vancouver reported that using virtual care in their CIED care processes decreased patient anxiety levels, improved communication of device information, and allows patients living in rural communities’ access to specialized care. Furthermore, they state that this integration has also presented challenges, such as changes to clinic workflow, compliance and usability by patients, and the need to develop effective education strategies when initiating monitoring with new patients. ^37, 38^ Similarly, Sparkes *et al*. conducted a usability study of a mailable, non-invasive cardiac testing kit to five participants, each with varying experience with technology. This specific application required independent set-up by patients, who were recorded doing so in a ‘think aloud’ manner, while completing 20 device set-up tasks, followed by an interview to elucidate their feelings about the process. While gender, age, and familiarity with technology seemed to influence the participants’ abilities to successfully set up these devices, they concluded that sending the kit by mail appeared to be an acceptable strategy. ^39^ However, given the small sample size, larger studies that could help to confirm these results are warranted. These could help to further elucidate the language that should be used in the instructions and the obstacles that should be anticipated in future deployments. Taken together, these studies show the importance of involving patients as stakeholders when designing virtual care technology and dissemination strategies. Also, if designed correctly, patients may not need to attend a preliminary face-to-face visit with their care providers before initiating a virtual care-based healthcare relationship. Established self-learning and orientation by video or walk-through modules is one way to empower patients in this regard.

Rush *et al*. from the University of British Columbia has done extensive research into the perspectives of physicians and patients living in rural areas concerning the use of virtual care for atrial fibrillation (AF), specifically. In a preliminary study, thematic analysis of semi-structured interviews with 14 patient and physician stakeholders revealed a high degree of variability in receptiveness to virtual care. This was reported to be a result of differences in past experiences with virtual care, in perceived adequacy of rural health services, and in perceived gaps in AF care. ^40^ In a follow-up study with over 10-fold more patients (n=116) from three rural communities, there were similar problems in managing AF. As it relates to virtual care, these researchers found that access to primary and cardiology care was a recurring challenge, and emergency department use was highly contentious but often the only option for accessing care. Primary care physicians were generally comfortable managing AF but varied in their reasoning for making referrals to specialists, often reserving them for complex situations to avoid the need for patient travel. The patients and providers supported a broad approach to virtual care of AF that is tailored to be inclusive of patients living in a rural demographic and thereby preserve the vital role of primary care physicians. ^41^

### Ischemic Heart Disease Interventions

Accurate and efficient interpretation of pre-hospital 12-lead electrocardiograms (ECGs) has shown to produce favourable patient outcomes as it reduces the delay to reperfusion through fibrinolysis or percutaneous coronary intervention (PCI) therapies. ^42^ Tanguay and colleagues reported on a virtual care-based STEMI detection program used throughout rural Quebec. This 7-year retrospective study included 728 patients with suspected STEMI who were transported by EMS. The transmission of an ECG every two minutes allowed for remote interpretation of abnormalities by a physician to discern between patients with a STEMI and those with a non-STEMI. The system enabled remote diagnosis of STEMI in 8.1% of patients during transport following an initial non-STEMI diagnosis. Thus, serial monitoring of these dynamic changes can allow for more rapid diversion to primary PCI facilities, potentially improving outcomes. ^42^ Such an approach could also lend itself well to machine learning decision support systems that improve triage or follow-up.

The American Heart Association guidelines recommend that the time between first medical contact and balloon inflation for STEMI patients should not exceed 90 minutes. ^43^ However, this is not realistic for many rural health systems, leading to increased morbidity and mortality for patients in these areas. The same group in Quebec also strove to address this issue by using virtual care services to reduce pre-hospital delays and make timely STEMI diagnoses. Over a similar 7-year time period, their retrospective data of 208 STEMI patients demonstrated that 14.9% were already on their way to a hospital with PCI capabilities, 75.0% were re-routed to a PCI centre, and 10.1% were directed to the nearest hospital. All patients but one arrived at the PCI centre within the guideline recommended 60-minute pre-hospital care interval. This study further illustrates that virtual care can help give timely access to PCI for rural populations that would not otherwise have access to this treatment. ^44^ Integrating virtual care with paramedic and emergency departments can further augment this capacity to deliver on-time care to rural patients. Despite this however, one could argue that truly remote patients will never be able to reach a PCI centre within the recommended time window, and thus these results may not be applicable.

Following an acute event of cardiac ischemia, cardiac rehabilitation programs, consisting of a combination of lifestyle and risk-factor management, can improve psychosocial outcomes and reduce both premature mortality and future cardiac events. Given that these programs are traditionally offered in large urban centres, geography is a common reason for decreased attendance and inaccessibility. The review by Lear summarizes key work done in Canada that leverages virtual care technologies for use in remote cardiac rehabilitation programs. ^45^ Lear *et al*. also conducted a pilot study followed by a scaled, randomized controlled trial; the trial was a 4-month virtual care-based program with a 1-year sustainability follow-up on exercise capacity and risk factor reduction compared with usual care. Only patients living in rural areas of British Columbia were enrolled, as this population was thought to benefit most from a virtual care program. For the primary outcome, after adjustment for the maximal time on the treadmill at baseline, age, sex, type 2 diabetes, and internet use for health information, participants in the treatment group had a greater increase in maximal time on the treadmill by 45.7 (95% CI, 1.04– 90.48) seconds compared with the control group during the 16 months (*P*=0.045). In unadjusted analyses, total cholesterol (−7.3%; *P*=0.026), low-density lipoprotein cholesterol (−11.9%; *P*=0.022), and dietary saturated fat (−1.4% kcal/d; *P*=0.018) were lower for the treatment group and required minimal resources with less than 8 hours of staff time per patient. Therefore, this model of care is cost efficient and readily sustainable. ^46^ A qualitative analysis of patient-physician discussions when using this virtual program has also been published, which suggests that simple chat sessions can be an effective alternative to in-person consultations, when necessary. ^47^

Pistawka *et al*. also conducted a virtual cardiac rehabilitation program in British Columbia that delivered health education to seven rural communities with a high prevalence of ischemic heart disease. The program was implemented over a 6-month period in partnership with local health authorities to ensure optimal uptake, ease of accessibility, and a streamlined referral process. This was followed by a six-week evaluation period, revealing robust patient interest, uptake, personal satisfaction, and improved self-efficacy, with over 90% of the study participants feeling more knowledgeable with better skills to manage their health. ^48^ This further emphasizes the importance of working with policy experts and procedural gatekeepers in health authorities to streamline innovation implementation and pilot processes.

### Hypertension Interventions

The prevalence of hypertension in Canada continues to rise and coordinated efforts to improve the treatment and control of hypertension are needed. ^49^ Despite the high prevalence, there is currently only one primary research article that investigated the use of virtual care for patients living in rural areas. Tobe *et al*. conducted the Diabetes Risk Evaluation and Management (DREAM)-global study to improve hypertension awareness, treatment, and management in Indigenous populations living in rural areas using virtual care. ^50-52^ They randomized 243 patients from six rural communities to receive text messages specific to hypertension management (active group) or general good health behaviours (passive group) and assessed for reductions in blood pressure over a one-year period. Despite an overall reduction in blood pressure, there was no difference in the blood pressure change between groups from baseline systolic 0.8 (95% CI −4.2 to 5.8 mm Hg) or diastolic −1.0 (95% CI −3.7 to 1.8 mm Hg, P = 0.5) blood pressure. Control was achieved in 37.5% (25.6%-49.4%, 95% CI) of the active group and 32.8% (20.6%-44.8%, 95% CI) of the passive group (difference in proportions −4.74% (−21.7% to 12.2%, 95% CI, P = 0.6). This suggests that virtual care can be effective for controlling blood pressure for Indigenous community members under the direction of community leadership. ^52^ In an editorial commentary about this study, Padwal suggests that passive virtual care interventions like text messages are unlikely to be effective and that more dynamic interventions such as tele-transmitting blood pressure devices are a better strategy. ^53^ Further cultural engagement with Indigenous communities is needed to have two-eyed seeing initiated with Indigenous ways of knowing to approaches and goals for CVD management. ^54^

### Cerebrovascular Disease Interventions

The phrase “time-is-tissue” is used by cardiologists and neurologists to emphasize that tissue is lost quickly as an infarction progresses without reperfusion; so emergent evaluation and rapid responsive therapy are required. ^55^ For this reason, virtual care is paramount to individuals affected by- and involved in the care of-patients with these conditions when geographically distant from healthcare centres specializing in stroke treatment. The Canadian Stroke Best Practice Guidelines state that virtual care services are a cost-effective tool to support health systems in closing the urban/rural and tertiary/primary care gap. ^56^

In their 2012 article, Dr. Hakim suggests that the delivery of thrombolysis with the help of virtual care is safe and achieves similar clinical outcomes as the same treatment delivered at stroke centres. He further suggests that a virtual care approach could be used to provide training, teaching, and therapy in the recovery phase for stroke patients living in remote regions. ^57^ Zimmer *et al*. discuss the training necessary to establish a stroke-focused virtual care system using a clinical toolkit in the Ontario Stroke System. The key components include clinical workflow algorithms, nursing/physician competencies, an education/training flowchart, protocols, technical readiness, evaluation tools, and resource contact information. Results from their study suggest that this training system facilitated efficiencies and standardization of practice to ensure optimal acute stroke management. ^58^ Taralson and colleagues also studied a similar training protocol applied in Alberta. They identified that an enthusiastic local leadership team, with strong physician support, and an equally supportive hub site stroke team are necessary for the implementation of successful remote stroke centres. Regular participation in mock code drills could also improve staff confidence and competency, leading to better patient outcomes. ^59^ This program established and developed evidence-based guidelines in order to implement virtual care across the continuum of stroke care. ^60^ Glasser and colleagues comment on the specific nursing competencies required to effectively use virtual care technology in this context, and stated that interpersonal communication and collaboration, and technology-related skills including the security of data and effective documentation of virtual visits are key areas to focus. ^61^ In patient centered care, where a degree of cognitive impairment is certain, facilitated care management using virtual care, appointment and instructional supports serves the individual and their informal care providers.

Khan *et al*. recounted one of the first Canadian virtual care experiences for acute stroke treatment in remote regions in which a critical care line was used by distant sites to connect to a physician with stroke-expertise at a hub site for patient examination and computed tomography (CT) scan assessment via a two-way video and audio link. Over a two-year period, the system was used 211 times, of which 45 patients (21%) were treated effectively without transfer. In this study, the number of transfers decreased 92.5% from 144 to 15 at one of the remote sites. These results suggest that virtual care could lead to a reduction in the number of patients requiring transfer to a tertiary care center benefiting quality of life and health economics. ^62^ Silver later provided an editorial commentary regarding the results—stating that the number of patients in the specific subgroups are too small to draw firm conclusions, but that the results suggest that virtual care could be non-inferior to in-person care in these settings. ^63^ Stroke quality indicators were later compared between patients treated remotely using this program and those who received care from an on-site stroke specialist, showing that they were indeed comparable. ^64^ Similar results were reported for the Ontario system. ^65^ The rationale for not yet nationalizing such a virtual care program remains unclear.

There is a need for CT-scans to assess cerebrovascular accidents to rule out brain haemorrhage risk prior to thrombolysis. This expensive equipment is not widely available in rural areas and remote healthcare sites, yet portable CT-scanners are brought into these rural sites and can be operated after minimal training. A portable CT-scanner at a remote site was implemented in the evaluation of patients who were not within timely reach of stroke experts. ^66^ Recently, a CT-equipped stroke ambulance was used to enable brain imaging at the patient’s location. This example of advanced paramedic technology suggests the potential for decentralizing care in remote areas to bring the “hospital to the patient.” While initial success has been documented, the question of cost-effectiveness, particularly in the rural setting, requires further study and is an important part of ongoing research studies. ^67^ Lastly, Mitchell *et al*. did a study about a promising radiology-focused emergency medicine program that would allow for urgent detection of brain hemorrhages, which could play an important role in the future of virtual care-based stroke care. ^68^

The disparities in CVD rates among Canadian Indigenous populations contribute to increased rates of stroke, and therefore some researchers have partnered with Indigenous organizations to work towards developing culturally appropriate services and support programs. Bodnar reported on projects undertaken within the Northwestern Ontario Regional Stroke Network centered around the development of educational materials and strategies, and virtual care-based initiatives designed to meet the unique needs of Indigenous peoples. ^69^ After stroke, extensive rehabilitation is often needed, involving physiotherapists, occupational (OT) and speech therapists, and social workers, many of whom are not accessible to remote populations, nor trained in cultural safety aligned with community values. French and colleagues reported on a multidisciplinary virtual care solution; over a five-month period, 10 consultations took place, with an average of 7.22 recommendations and referrals per visit (range 4-10). These consultations focused on mobility, education, resuming roles, and community participation. The experience was rated good to excellent by both patients and clinicians for 95.2% (SD=0.44) of the consultations. However, audio quality was rated as poor to fair by the patients in 75.0% (SD=0.82) of visits. All participants reported as willing to recommend the system to others. As is a common theme in this review, healthcare, community, and virtual care partnerships were critical to the success of this program. ^70^ Linkewich and colleagues reported on an analogous virtual care tool used to conduct OT home safety assessments for a similar patient population. The OTs who participated reported confidence in making practice recommendations using the program, however, a comment about needing to “sense” the client in a different way was repeatedly made, suggesting that reduced non-verbal body language could be a disadvantage of the tool. The client and all OTs were willing to recommend the virtual assessment system. Audio quality was also reported to be problematic, as in the previous study. ^71^ Technology based experiential quality is a surmountable issue and can be overcome through cell-tower and emerging satellite high-speed connectivity. These findings also emphasize the goal of virtual care should not be to eliminate in-person systems but rather focus them and integrate with it for productivity, efficiency, and patient outcome or experience benefit.

Moving On after Stroke (MOST®) is a group-based, self-management program for stroke survivors and their caregivers consisting of information sharing, facilitated discussions, goal-14 setting, and physical exercise. Taylor *et al*. simultaneously delivered the program to local participants onsite in Thunder Bay, Ontario and via virtual care to participants living in small, remote communities. Their objective was to explore the experiences of remote participants and their perceptions regarding factors that enable or limit their participation, and to obtain suggestions for enhancing delivery. As such, we can appreciate the preventative health benefit of virtual care. Interviews, followed by thematic analysis, with 19 participants revealed that there was value in not having to travel long distances for appointments. Many patients reported “feeling as if they were in the same room” but some patients acknowledged that there were limitations such as a loss of subtleties in communication. Some facilitators found it difficult to discern whether the exercise routines were appropriate for the participants. A preliminary face-to-face meeting and enhanced local support were found to improve the experience. ^72^ Patient compliance and preventative health can be a combined consideration in healthcare and environmental sustainability—respect the person and the planet by reducing the distance between people.

Whelan *et al*. reported results from the Saskatchewan Cerebrovascular Center virtual care study regarding consults and post-operative follow up visits. Over a two-year period, 184 patients who were surveyed following virtual visits saved an average of 243 kilometres, which was a cumulative total of 43, 948 kilometres saved and an average patient-perceived financial savings of $445 CAD per visit. All but one patient (94%) reported being satisfied. However, all patients were willing to use the system again. ^73^ Sakakibara *et al*. reported findings of a similar program based in Vancouver, whereby 102 patients (of which 37.2% were rural) completed a survey about their experiences using a system designed specifically for low-cost consumer technologies. While they found that the delivery of rehabilitation services using these technologies was both feasible and desirable by many stroke survivors, 71% of respondents believed their quality of care would be less than in-person rehabilitation. Therefore, the authors concluded that virtual care should augment and not replace in-person rehabilitation, but in cases where in-person rehabilitation is neither accessible nor possible, then virtual care is an acceptable substitution. ^74^ Lastly, Appireddy, *et al*. reported results from a 6-month study using virtual care for post-stroke follow-up care throughout the Kingston Health Sciences Centre in Ontario. This study included 75 participants (of which 39% were rural) and they found that: 1) There was a statistically significant shorter wait-time for a virtual appointment compared to in-person (median 60 vs 78 days). 2) The virtual care visits were also shorter in duration, taking on an average of 10 minutes to deliver follow-up care with a high degree of patient satisfaction versus an average of 90 minutes for in-person care. 3) On average, the total time saved by patients per visit was 80 minutes, of which 44 minutes was travel time. 4) Travel distance avoided by the patients was an average of 30.1 km. 5) The estimated total out-of-pocket cost savings for patients per visit was $52.83. 6) The estimated savings (opportunity cost for in-person outpatient care) for their entire virtual care pilot project was $23,832 - $28,584 CAD. The patient satisfaction with the virtual system was good compared with their prior personal experience with in-person outpatient care. ^75^ A point worthy of exploration is the discrepancy in patient cost-savings mentioned in this study as compared with the figures cited previously of $445 and $500 CAD. The fact that this study contained only 39% rural patients, whereas the other two studies were entirely composed of rural patients may indicate that much of the cost-savings are likely related to food, parking, accommodation, and not simply mileage. This discrepancy may also reflect the fact that rural Saskatchewan and Alberta have greater cost to distance travel ratios than Ontario, or perhaps the cost discrepancy is due to lack of inflation adjustment. Incorporating health economic analyses to virtual care pilots, implementation, monitoring should be emphasized. Health value demonstration initiatives are ideally well powered and longitudinal partnerships between academic institutes and health authorities.

## Discussion

To our knowledge, this is the first review of recent publications that examines the use of virtual care technology seeking to benefit patients with CVD who are living in rural Canadian locations. We have attempted to understand the effectiveness of the various interventions, which have generally had positive effects among the patient population of interest. The specific subjective benefits of virtual care include reduced hospitalization and readmissions, improved mortality rates, increased cost-effectiveness for patients and hospitals, improved quality of life for patients, and an increase in self-management of CVD. Further benefit is derived through better disease prevention and increased patient investment in their health span. Virtual care is not a panacea for all rural medicine needs. However, it is clear that value is being extracted, predominantly by three Canadian provinces (BC, ON, and QC), which is surprising since these provinces have the greatest urban concentrations of people. ^76^ These provinces also account for the majority of research pertaining to rural medicine in general, which indicates potential inequities in medical research contrary to the objectives of the Canada Health Act. Remarkably, the Western Prairies, Territories, and the Atlantic provinces (with seemingly the most need and the most to gain from virtual care innovations) are not well represented in this subsection of the literature. In the Canadian context, this puts all provinces at a disadvantage by being over-reliant on health transfers from the federal government instead of leadership by strength. Establishing an online, virtual care institute amongst low population density provinces might be possible, yet most supercluster and centers of excellence are concentrated in dense urban zones. Exploring this potential inequity from a legal, social and humanities perspective should be of interest to researchers moving forward.

While still largely unadopted on a national level, the data indicates that systems in place for acute pathologies like stroke and STEMI are the most well-established and far-reaching, while systems focused on more chronic subtypes of CVD like HF, hypertension, and arrhythmias, are lagging and largely remain in the pilot phase. This is an important point to highlight, as the management of chronic disease is likely to have the greatest impact. Studies performed within government by health departments or industry in the form of white papers, were inaccessible for review. As a result, very few randomized control trials were identified and much of the work in this area is only being shared in conference proceedings. In the wake of the COVID-19 pandemic, this dynamic is likely to rapidly evolve given the expedited policy changes surrounding the use of virtual care technologies. ^77^ Innovative solutions like the ones discussed here are now of critical importance given their ability to reduce contact between healthcare providers and vulnerable patient populations.

Overall, there is an interest to support virtual care among the stakeholders investigated in this study, despite some perceived barriers when compared to in-person approaches to healthcare. Despite this enthusiasm, there is still insufficient evidence to objectively support the outcomes, health economics, physician/patient-satisfaction, and privacy concerns due to a lack of trials and peer-reviewed evidence at this time. CVD risk factor management using virtual care needs peer reviewed study in the Canadian context to build an evidence base that informs policy development. Such studies should emphasize the rationale, define the intended benefit and the comparator arm, and address cost-effectiveness or any barriers to implementation. Until that time, it remains presumed but uncertain that virtual care in Canada can assist in the prevention of CVD, when compared with more specified conditions requiring interventional support.

Despite the fact that the collective studies lacked the desired scientific rigour, or are of limited meta-data, the qualitative insights provide valuable recommendations for researchers planning to deploy virtual care technologies in the future. These have been summarized in Figures 3 and 4. Briefly, a common point of note was the need to involve patients as stakeholders during the planning of virtual care distribution strategies. Kim and colleagues provide insightful commentary as to how best to involve patients. ^78^ Another was the need to clarify the roles and responsibilities of all members of the healthcare team related to the virtual care system, specifically stressing who will be responsible for the training of patients and informal care providers. This may represent an obstacle best overcome by online education videos or other patient assistive products and services. Role clarification is needed between the payer and the provider for services and should be explored in future studies. Canada remains predominantly a single/public-paying system and is slow to integrate innovations predominantly developed by the private sector. Establishing *a priori* criteria to be met by the private sector to deliver virtual care would be welcomed, as the preferences of payers are a critical policy consideration for health authorities. Establishing victory conditions (that is, clear target endpoints), would avoid any further languishing of incorporating healthcare technology innovation that is greatly needed, and so as to be prepared for crisis rather than react to it in the example of implementing virtual care during a pandemic.

**Figure 3.**
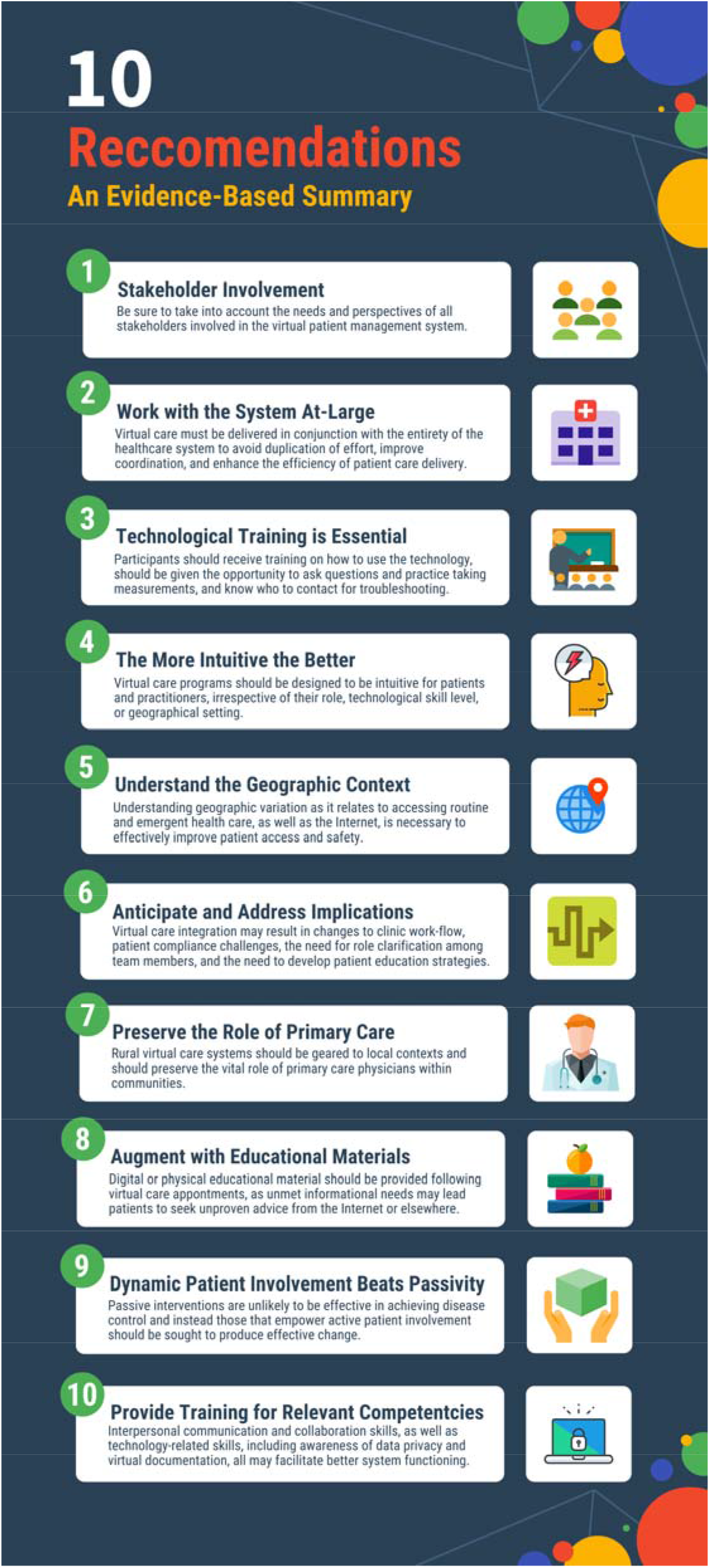
Summary of virtual care recommendations.

**Figure 4.**
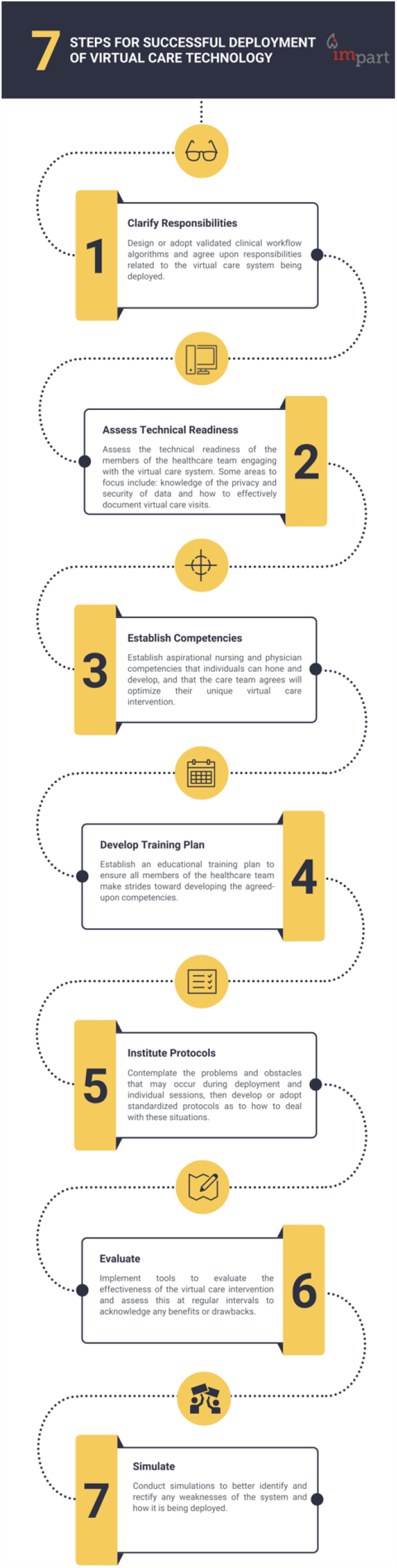
Summary of practical objectives for future virtual care programs.

The current disparities in health metrics suggest that patients in rural areas, particularly Indigenous nations, desire greater diversity in healthcare delivery, desire to contribute culturally and require specific accommodations. In addition to virtual care services, advanced care paramedics, mobile care units, increased scope of practice for rural pharmacists, augmented extramural nursing and home care options, and physician assistants, could all contribute to rural healthcare teams. Many of these auxiliary topics, in addition to the challenges of policy and privacy, were not well addressed in these studies and should be the subjects of future research and policy development. Similarly, little in the way of economic analyses were reported, nor is there a consensus for adopting cost-saving or cost-deferring innovations or establishment of time-dependent value-thresholds. Those studies that did report on the cost savings of the intervention averaged ∼$475 CAD per appointment for patients living in rural communities. Using a reported rural Canadian population of 19%, data showing ∼500,000 HF patients, and an estimated 10 HF-related healthcare appointments per year per patient, equating to $451Million CAD in lost opportunity, illustrating that virtual care tools could significantly reduce out-of-pocket expenses for patients. ^79, 80^ To further illustrate, we will assume that 60% of all in-person visits could be performed using virtual care tools and systems, as has been the low estimate during the COVID-19 pandemic. The Commonwealth Fund reports that the average annual physician visits per capita in Canada in 2017 was 6.8. ^81^ For a current population of 37.59M, and an opportunity cost ranging between $31-$500 CAD per visit, this extrapolates to potential cost savings of ∼$4.8 - $77 billion CAD. This is to say nothing with respect to the cost savings for the healthcare system itself, which would likely be enough to recoup the costs of purchasing and implementing a virtual care system within a short period of time. Industry partnered health value demonstration initiatives dedicated to regions with a high need can accelerate access to this opportunity by engaging with academic institutes and health authorities in tri-lateral objectives to patient-centered care.

### Study Limitations and Strengths

This systematic review has limitations. We evaluated published articles written in English from three relevant databases, therefore, virtual care interventions described in other languages or those not peer reviewed but communicated internally within government health departments were likely missed. Additional search results might have been found in other databases and sources (eg, grey literature, books) and were therefore not included in this review. We also did not evaluate articles that were published prior to 2010, as we decided that 10 years was a sufficient amount of time to gather material germane to our objectives in a country with a paucity of standard virtual care. Further, only a single reviewer screened the included studies, therefore unconscious selection bias may be at play, however studies show that this approach still represents an appropriate methodological shortcut. ^82^ Strengths of this review include the use of an experienced librarian for the development of the search strategy and adherence to the PRISMA guidelines.

### Future Directions

Many important areas relating to virtual care were not well addressed and require further study. These include: 1) the role of the public and private sectors in evaluating, disseminating, and overseeing the use of virtual care technology, 2) appropriate remuneration rates for using virtual care technology, 3) incentivizing *a priori* criteria for using/adopting virtual care technology, and 4) preparing future healthcare providers to incorporate virtual care technology in cardiovascular practices, such as providing training in undergraduate medical education programs and providing continuing medical education credits.

## Conclusion

In sum, it is clear that efforts are being made to ensure equitable care of patients diagnosed with all subtypes of CVD, although future studies targeted in the realm of hypertension management among rural Canadian populations are greatly needed. The systems for acute pathologies seem to be the most well-established, while those focused on chronic cardiac conditions remain in the pilot phase. The evidence presented here also indicates potential for long-term healthcare savings with virtual care solutions. Given the themes identified in this review that virtual care technologies are efficacious, well-accepted, and economical, we feel that these systems should be adopted and better integrated into clinical cardiology practices in order to care for patients diagnosed with CVD living in rural areas of Canada. Funding and partner agencies should prioritize high-risk high-reward regions with considerable rural populations to maximize early returns on investments in virtual care.

## Data Availability

No datasets were generated or analyzed during the current study.

## Acknowledgments

The authors would like to acknowledge the help of Jackie Phinney (Librarian, W.K. Kellogg Health Sciences Library, Dalhousie University) who assisted in the development of the literature search strategy.

## Appendix

